# Effects of a Remote Mental and Physical Practice Intervention on Freezing of Gait in People with Parkinson’s Disease: a Single-Blind Randomized Controlled Trial Protocol

**DOI:** 10.1101/2025.04.28.25326452

**Authors:** Paloma Rodrigues da Silva, Karina Yumi Tashima Honda, Marina Rigolin Pikel, Anelise Rodrigues dos Santos, Maria Elisa Pimentel Piemonte

## Abstract

**Background:** Freezing of gait (FOG) is a common motor symptom in Parkinson’s disease (PD), increasing the risk of falls and reducing independence and quality of life. Effective management combines pharmacological treatment and, when necessary, surgical intervention with targeted strategies aimed at improving gait through compensatory brain networks. Mental Practice (MP), which consists of mentally rehearsing movements, has been studied as a possible strategy to facilitate gait through goal-directed motor control. However, there is limited evidence regarding its therapeutic effects on FOG. This study aims to compare the effects of a novel intervention combining MP based on Dynamic Neuro-Cognitive Imagery (DNI) and Physical Practice (PP) with a control intervention involving PP only, both delivered remotely on the severity of FOG episodes in individuals with PD.

**Methods:** A single-blind, controlled, and randomized trial is proposed. People with idiopathic PD who answered positively to the first question of New Freezing of Gait Questionnaire (NFOG-Q) will be randomly allocated into the Experimental Group (EG) and Control Group (CG). Both groups will undergo 10 remote intervention sessions lasting 45 - 60 minutes with identical volume and intensity, differing only in the MP components. The EG will perform sessions of MP combined with PP, while the CG will perform sessions of PP and stretching. They will be assessed before intervention (BI), after intervention (AI), and 30 days after the intervention, as a follow-up (FU). Primary outcomes included the time to complete the Rapid Turn Test, and the Percentage of time spent with FOG during the test (%FOG). Secondary outcomes included the NFOG-Q, the Movement Disorders Society – Unified Parkinson’s Disease Rating Scale (MDS-UPDRS), the Parkinson’s Disease Questionnaire-39 (PDQ-39), and the Telephone Montreal Cognitive Assessment (T-MoCA). A repeated measures ANOVA will be used for statistical analysis.

**Discussion:** The main hypothesis is that both interventions, which are similar in terms of volume, intensity, and progression of complexity, will positively affect the FOG severity. However, experimental intervention combining MP and PP is expected to produce better outcomes than the control intervention. If this hypothesis is confirmed, the study could significantly impact the treatment of people with PD by providing a low-cost and widely accessible option.

**Administrative information:** Note: The numbers in curly bracelets in this protocol refer to SPIRIT checklist item numbers. The order of the items has been modified to group similar items.

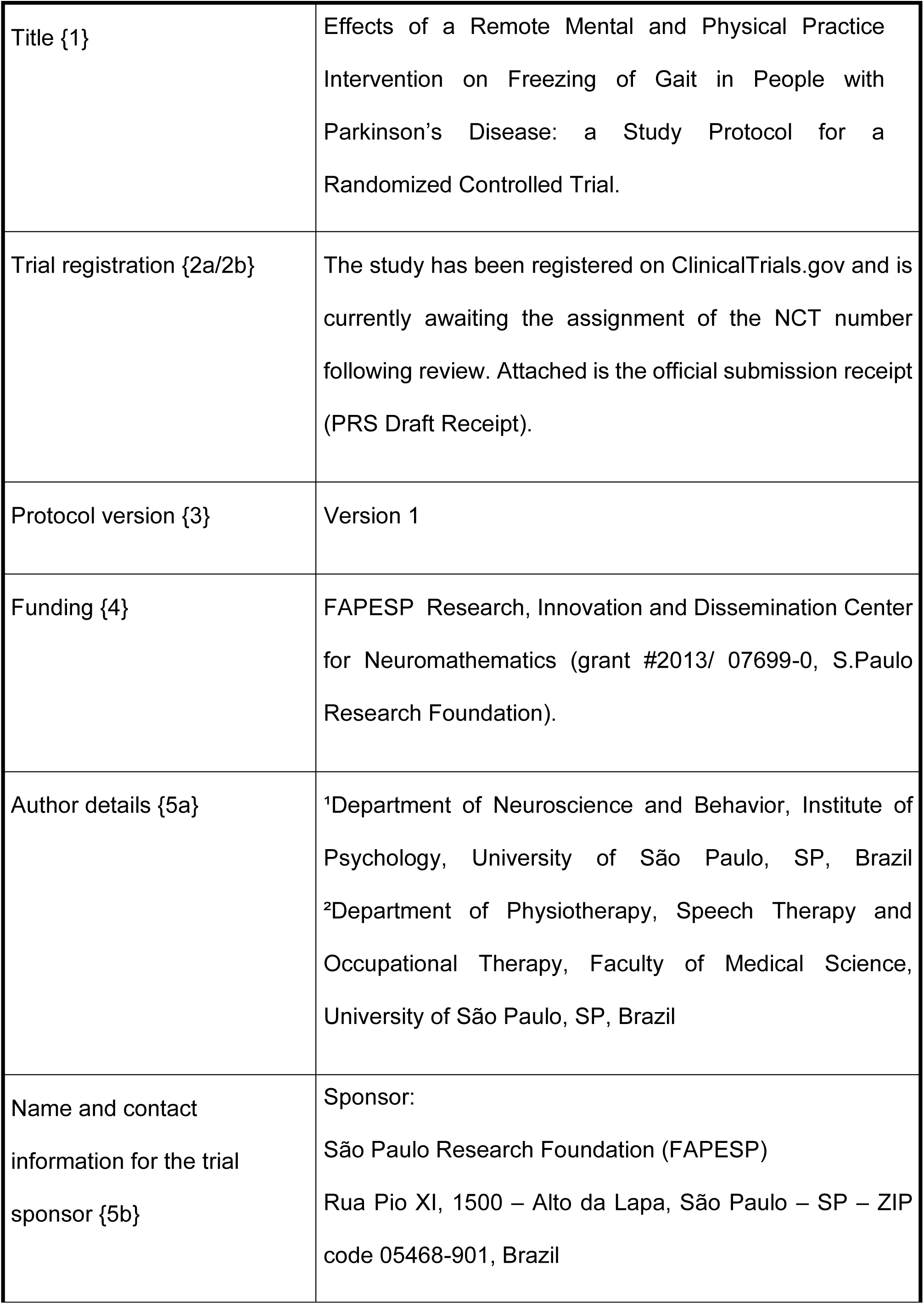

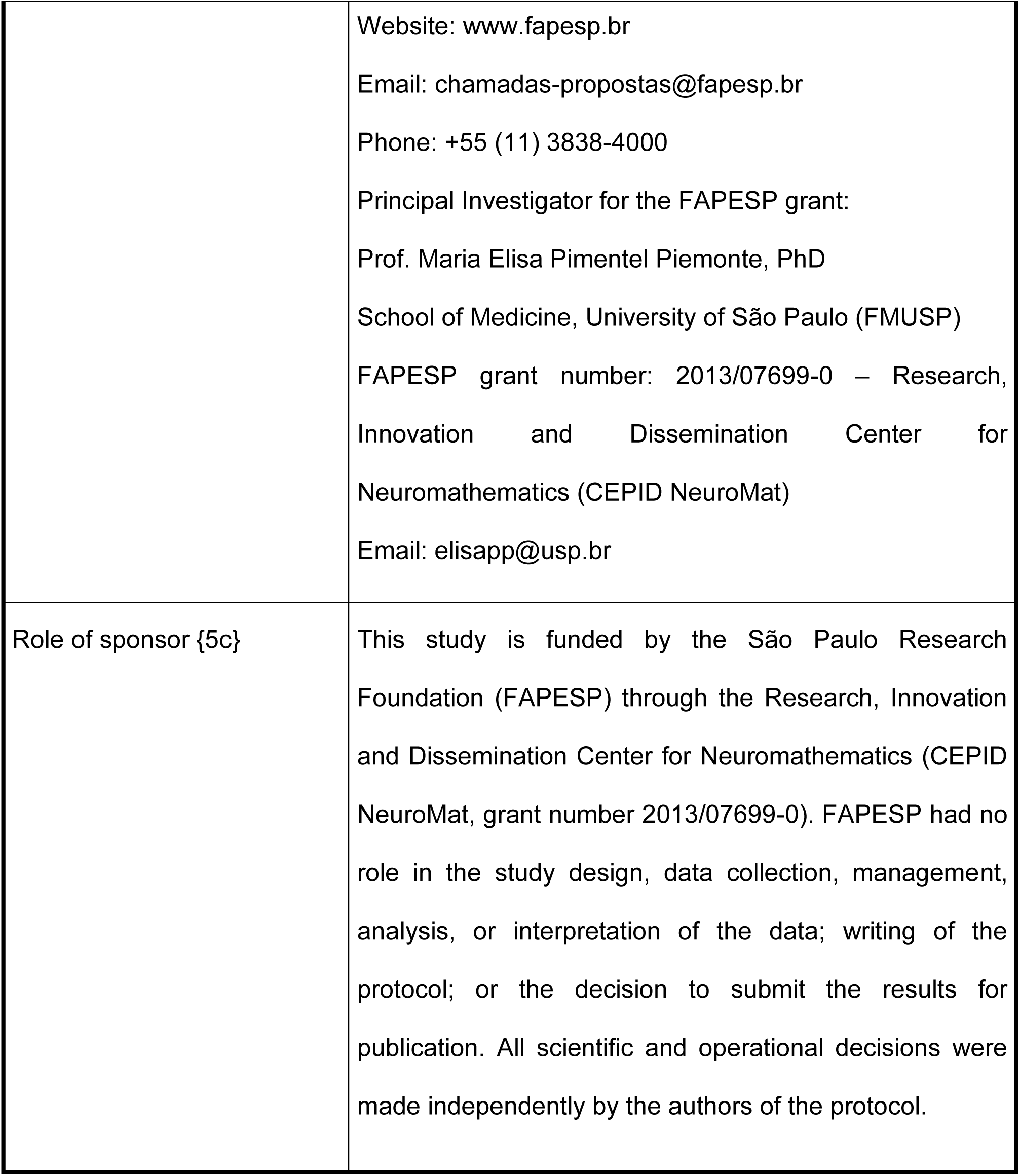

## Introduction

### Background and rationale {6a}

Changes in gait are one of the most debilitating motor symptoms of people with Parkinson’s Disease (PD), causing a high negative impact on autonomy, independence, and quality of life [1,2]. Gait is a complex process that involves automatic, emotional and voluntary control, in addition to demanding cognitive resources [3]. Gait impairments in PD can be divided into two main categories: those that are continuously present and those that occur episodically. Continuous disturbances include reduced step length and height, decreased gait speed, increased gait variability, stooped posture, and asymmetrically reduced arm swing. Episodic disturbances include the excessiveness of the festination and freezing of gait (FOG) [1].

FOG is defined as “a brief, episodic absence or a marked reduction in forward progression of the feet, despite the intention to walk”, associated with the sensation that “the feet are glued to the floor” [4,5]. This symptom is associated with a deterioration in quality of life, limiting their mobility, increasing the risk of falls, and contributing to social isolation [6].

Due to its significant impact on patient functioning and limited response to existing therapies, FOG has been prioritized as a critical research focus to enhance understanding and develop more effective treatments [7].

The FOG pathophysiology involves a complex dysfunctional integration of motor, cognitive, and limbic networks and is not completely understood. Currently the two key dysfunctions are associated with FOG pathophysiology: (1) excessive inhibitory basal ganglia outflow due to dopaminergic degeneration in the nigrostriatal pathway which disrupts brain stem locomotor centers causing the de-automatizing the gait control, and (2) lack to activate the compensatory network integration which is crucial for maintaining attention and integration between cognitive (prefrontal cortex), sensory (parietal cortex), and motor networks during gait challenges. This limits the ability to compensate for the loss of gait automaticity, which is essential for avoiding FOG in complex situations [8].

Several evidence confirms the association between disruption in automatic gait control and FOG in PD. People with PD and FOG showed a significantly more severe loss of automatic gait control than those with PD who do not have FOG [1] and increased dual-task cost during gait, even after controlling for disease severity and global cognition [9]. Furthermore, recent findings from resting-state fMRI analyses suggest that the effectiveness of cueing strategies to alleviate FOG depend on the engagement of compensatory executive-attentional brain networks [10]. These findings suggest that people with FOG rely more heavily on attention to maintain gait performance, reflecting reduced automaticity and increased need for top-down modulation.

Based on this FOG pathophysiology model, the more recent expert recommendations pointed two key to optimal treatment: (1) reduction of excessive inhibitory outflow of the basal ganglia through optimization of dopaminergic therapy and advanced interventions, including infusion therapies and Deep Brain Stimulation; and (2) facilitation of compensatory networks, including physical training and fall prevention, and interventions to engage compensatory cortical networks by allocating more attention to gait, thereby reducing reliance on the automatic gait control system [8].

Two recent systematic reviews have shown a growing body of evidence on exercise and rehabilitation interventions for FOG in PD. Gilat et al. (2021) demonstrated a moderate overall effect of exercise-based interventions on self-reported FOG severity, particularly for targeted interventions such as cueing strategies and cognitive-motor training, whereas generic exercises such as conventional physiotherapy showed no significant effects [11]. Goh et al. (2023) also identified a small but significant reduction in FOG severity; however, subgroup analyses did not confirm superiority of specific modalities [12]. Taken together, these reviews suggest that while certain exercise-based interventions may offer modest benefits for FOG, their long-term effectiveness and specificity remain inconclusive, reinforcing the need for developing more effective interventions.

Among the interventions to facilitate compensatory networks, cognitive-motor training that includes observational training, explicit or internal cueing, dual-task training and mental motor imagery, has been considered a powerful strategy to activate higher-order cortical networks to minimize the impaired automatic gait control in PD. Action observation training (AOT), a kind of mental practice (MP) guided by visual information, is effective in reducing FOG and inducing functional reorganization in motor and fronto-parietal attentional networks correlated with FOG improvement [13,14].

MP consists of mental rehearsal of movement, experienced from a first- or third-person perspective, without actual physical execution, with the aim of supporting motor learning and improving performance, particularly in people with neurological conditions [15]. It activates brain regions similarly involved in Physical Practice (PP), such as the premotor cortex, supplementary motor area, cingulate cortex, parietal cortex, basal ganglia, and cerebellum [15,16,17]. The mechanisms underlying MP involve both neuropsychological and neurophysiological components. The first one, enhances aspects such as task segmentation, attentional focus, and the development of alternative movement strategies. The second one promotes neural adaptations in motor planning networks within the central nervous system [18]. These effects are often explained through an ideomotor framework, which posits that anticipating sensory outcomes plays a critical role in motor learning. MP, as a multisensory experience, recruits perceptual systems to simulate action, allowing for error detection and refinement of internal representations. This makes it a powerful tool for improving performance and skill acquisition [19].

MP is thus considered a form of internal training whose effectiveness is closely linked to the engagement of internal models - particularly forward models - that simulate the sensory consequences of imagined actions. These models generate predictions based on efference copies of motor commands and are shaped by prior sensorimotor experiences. The relative weighting of visual, auditory, and proprioceptive inputs depends on the task demands, influencing how the imagery is constructed. Rather than being a passive cognitive process, MP dynamically mirrors real-time planning and feedback mechanisms, providing an internal feedback loop that facilitates motor adaptation even without overt movement [20].

Neuroimaging evidence suggests that MP may activate compensatory cortical networks involved in overcoming the loss of automatic gait control in people with PD and FOG. Task-based Functional Magnetic Resonance Imaging (fMRI) study demonstrates that, during imagined gait tasks, people with FOG exhibit increased activation in parieto-occipital and frontoparietal cortical areas, alongside reduced connectivity within subcortical locomotor circuits. The enhanced engagement of higher-order cognitive and visuospatial regions during MP reflects a compensatory strategy aimed at modulating motor output through alternative cortical pathways. Therefore, interventions based on MP may facilitate the reactivation of motor plans and support top-down control mechanisms, making them a promising non-pharmacological approach to address FOG in PD [21].

Although people with Parkinson’s disease (PD) retain the ability to generate detailed and vivid mental images [22,23], the evidence regarding the efficacy of MP for improving gait performance remains limited. While an increasing body of literature supports the incorporation of MP as an adjunctive strategy in the rehabilitation of people with PD, substantial methodological limitations, small sample sizes, and heterogeneity across study designs and outcomes continue to challenge the generalizability and strength of current findings. A scoping review including 53 studies report that MP interventions - especially when focused on gait and balance - can yield significant improvements in key functional outcomes such as walking speed, Timed Up and Go (TUG), step length, and dynamic balance. Notably, protocols combining MP with physical practice or action observation therapy induced measurable changes in both clinical metrics and neuroplasticity, including enhanced activity in brain regions involved in gait control. However, the authors emphasize the limited number of high-quality randomized controlled trials, the lack of standardized MP protocols, and the absence of studies targeting upper limb function or speech. While preliminary evidence indicates that MP may facilitate compensatory cortical engagement and improve motor planning in patients with FOG, larger and better-controlled studies are needed to confirm its therapeutic value and define optimal implementation in PD rehabilitation [24].

Although previous studies have shown positive effects using observational training that involves visual MP (third person) in PD [25,26], evidence from fMRI showed that kinesthetic MP is more efficient in recruiting parietal and frontal regions of the brain, which are critical for sensorimotor integration, in comparison to visual MP, that primarily activated areas related to visual-spatial processing (27). The evidence from a randomized clinical trial comparing neurofeedback-guided with kinesthetic and visual imagery in people with PD showed that, while both groups improved in motor function, only the group using kinesthetic MP showed a correlation between neurofeedback regulation of frontal cortex connectivity and functional gains. Additionally, participants in the kinesthetic imagery group reported increased body awareness and perceived improvements in balance, stride symmetry, and reduced FOG episodes (28). These findings suggest that first-person kinesthetic imagery may promote superior therapeutic results in PD rehabilitation.

Among the emerging approaches, Dynamic Neuro-Cognitive Imagery (DNI) may be an advantageous therapeutic option to improve key mechanisms implicated in motor planning and execution in PD. DNI is a structured, multisensory, imagery-based training approach designed to enhance motor performance through anatomical embodiment and kinesthetic awareness [29]. Preliminary evidence indicates that DNI may promote the internalization of effective movement strategies and cueing mechanisms, improving functional motor outcomes related to balance and lower limb coordination, suggesting potential benefits for gait rehabilitation in people with PD [18]. However, only two studies have investigated the effects of DNI in people with PD. A randomized controlled trial with 20 participants with PD showed significant improvements in mental imagery ability, disease severity, and motor and cognitive function for the participants who received two weeks of training based on DNI compared to participants who received the control intervention (a home-exercise video program) [30]. A second randomized clinical trial with 20 participants with PD showed that only participants who received DNI intervention demonstrated significant improvements in pelvic schema and the accuracy of graphic-metric representation after a two-weeks intervention [31].

Remote healthcare methods are valuable tools linked to technological advances, crucial for managing neurological and movement disorders like PD. These individuals often require long-term care and experience physical and cognitive limitations, making visits to specialized centers. Additionally, issues like lack of transportation and limited financial resources can worsen the situation [32]. Telerehabilitation, a modality of remote healthcare, is a promising way to deliver rehabilitation services remotely [33] and the use of this tool can contribute to improvements in motor performance and quality of life for people with PD, positively impacting mobility, cognition, and social support [34,35]. Therefore, this type of resource emerged as an alternative way to manage and supervise home exercise programs [32].

Although preliminary evidence suggests that MP may improve gait performance in people with PD, to our best knowledge, no randomized controlled clinical trials have investigated specifically the efficacy of a remotely delivered MP on FOG severity. To address this gap, the present study proposes a novel intervention combining MP based on DNI principles with PP. This approach aims to provide a low-cost, accessible, and scalable solution for reducing FOG episodes.

Thus, the main objective of the present study is to propose a protocol for a single-blinded, controlled, and randomized clinical trial to assess the effects of this innovative remote intervention compared to a control intervention involving only PP on FOG severity in people with PD.

### Objectives {7}

The primary objective of this study is to compare the effects of a novel intervention combining MP based on DNI and PP with a control intervention (CI) involving PP only, both delivered remotely on the severity of FOG episodes in individuals with PD.

Secondary objectives include evaluating the effects of the interventions motor aspects of daily living, quality of life, and cognitive function.

To achieve these objectives, a single-blind, controlled and randomized trial is proposed in which the effects of the experimental intervention (EI) will be compared with those of a CI, identical in volume and intensity, apart from the MP content.

The main hypothesis is that both interventions, which are similar in terms of volume, intensity, and progression of complexity, will positively affect the FOG severity. However, experimental intervention combining MP and PP is expected to produce better outcomes than the control intervention. If this hypothesis is confirmed, the study could significantly impact the treatment of people with PD by providing a low-cost and widely accessible option.

### Trial design {8}

A single-blind, controlled and randomized trial design according to the Standard Protocol Items: Recommendations for Interventional Trials guidelines (SPIRIT).

The study was approved by the Research Ethics Committee of under protocol number 7.253.458. All procedures will comply with the ethical standards of the 1964 Helsinki Declaration and its later amendments, as well as with the Brazilian National Health Council Resolution 466/2012.

## Methods: Participants, interventions and outcomes

### Study setting {9}

Participants will be recruited consecutively from the contacts of the AMPARO network (www.amparo.numec.prp.usp.br) by a non-probability sampling method. The clinical information will be obtained from the healthcare system records where patients receive medical care for PD. The records will be no older than six months to ensure that the information accurately reflects the participants’ current condition and to avoid outdated information. This information will be subsequently verified with participants and their families. Details of dopaminergic treatment, including drug type and daily dose, will be recorded to calculate the levodopa equivalent daily dose (LEDD).

### Eligibility criteria {10}

People with PD following criteria: medical diagnosis of idiopathic PD based on the London Brain Bank criteria [36], use of dopaminergic medication, positive response to the first question of the New Freezing of Gait Questionnaire (NFOG-Q) [37], Able to walk independently at home, with access to internet and video call device, and agree to participate in the study.

Exclusion criteria include the presentation of other neurological disorders, severe cardiovascular and/or respiratory alterations, uncorrected visual and/or auditory alterations, cognitive deficits that prevent the understanding of verbal instructions indicated in the protocol, detectable through the Telephone Montreal Cognitive Assessment (T-MoCA), with a score lower than 12 [38], and those who are unable to perform motor imagery during the administration of the Kinesthetic and Visual Imagery Questionnaire - 20 (KVIQ-20) scoring equal to or below 20 [39], participation in concurrent physiotherapy programs specifically targeting gait or FOG; changes in antiparkinsonian medication within 30 days prior to enrollment; and unstable medical or psychiatric conditions that could interfere with study participation.

### Who will take informed consent? {26a}

The selected subjects will be invited via internet and/or telephone to participate in the study. They will be informed about the research objectives and procedures and after showing interest they will be invited for an initial assessment with a scheduled date and time. At this time, they will be informed about the study again and receive more detailed information. Those who agree to participate will sign an informed consent form sent online, including options for the participant to take a screenshot, download, and/or print, along with guidance to keep a copy in their possession.

### Additional consent provisions for collection and use of participant data and biological specimens {26b}

Not applicable, data collection for ancillary studies will not be required.

### Interventions

#### Explanation for the choice of comparators {6b}

The effects of the EI will be compared with those of a CI, identical in volume and intensity, except for the MP content.

#### Intervention description {11a}

All assessment, reassessment, and intervention procedures will be conducted remotely via real-time video calls. Both intervention, experimental and control, will consist of 10 training sessions, each lasting 45-60 minutes. Sessions will be scheduled twice per week during the first four weeks and once per week during the final two weeks, totaling six weeks of intervention.

All sessions will be conducted synchronously by physiotherapists who will receive prior training to ensure full adherence to all procedures and protocols specific to both intervention arms (experimental and control). This prior training will be delivered by two in-person sessions by the principal investigator. The training will cover all components of both protocols, including session structure, verbal instructions, and remote delivery procedures. To ensure consistency and competence, physiotherapists will be evaluated through supervised simulations with volunteer patients by the principal investigator. Only physiotherapists who demonstrate full proficiency and uniformity in delivering the intervention will be considered prepared to begin training sessions with study participants.

### Orientation for adequation of home environment

Before the beginning of assessments and interventions, physiotherapists will provide to participants and, when applicable, their family members or caregivers, detailed guidance on how to prepare the home environment to ensure safety, comfort, and technical adequacy for the remote sessions. These orientations will include instructions on how to establish a stable and secure internet connection and how to position the device to allow for clear visual and auditory contact between the physiotherapist and the participant, ensuring effective communication throughout the session. Additionally, they will provide guidance on preparing the home environment to ensure a proper and safe training space. This will include guidance on selecting a comfortable and stable chair for participants to follow procedures in a seated position. For exercises performed in a standing position, they will provide guidance for organizing a clear space of at least two meters in length, free of rugs or obstacles that could pose a fall risk, with easy access to a stable support surface (e.g., wall or table) to be used for support in case of an unbalanced situation.

Participants will be instructed to wear safe, non-slip footwear and avoid loose clothing or accessories that could interfere with movement. In addition, they will be introduced to the materials required for the PP, such as toilet paper rolls, a 500 ml plastic water bottle, a broomstick, and a half-filled glass of water, and will be instructed to keep these materials available for use during the sessions.

Caregiver involvement during this preparatory phase will be encouraged to facilitate adherence to the instructions and ensure a safe environment throughout the intervention.

### Education on FOG (First session)

In the beginning of the first session, prior to the initiation of training, all participants will receive a structured educational module delivered by the physiotherapists. This session aimed to provide a comprehensive understanding of FOG and its implications for decline in independence and quality of life. Participants will be educated about the definition of FOG, its underlying pathophysiological mechanisms, and its relationship to the progressive gait disturbances related to PD. It will be emphasized that while these motor impairments are largely irreversible due to neurodegeneration, their functional impact can be minimized by compensatory strategies. The discussion will also include common internal and external FOG triggers, such as environmental constraints and emotional factors like anxiety, as well as strategies to identify and anticipate FOG episodes. Explanations will be supported by illustrative figures to facilitate understanding of the concepts discussed. It is important to highlight that the session will be conducted in an informal and conversational manner, allowing participants to share their own experiences with FOG, express concerns, and ask questions freely. The participants will be encouraged to request additional clarification at any point during the intervention.

In addition, after the educational content on FOG is presented, familiarization will be conducted with the MP for the Experimental Group (EG) - with an emphasis on ankle mobility and postural adjustments - and with the stretching exercises for the Control Group (CG).

### Experimental Intervention

The remaining nine sessions of the EI will consist of two blocks of MP and two blocks of PP, each lasting 10 minutes, totaling 40 minutes of training per session (20 minutes of MP and 20 minutes of PP).

The PP and MP blocks will be structured to provide progressive challenges in terms of complexity for gait control. Over the course of ten sessions, the content will be presented in a fixed and gradual sequence, ensuring that all blocks are introduced and developed over time.

#### Mental Practice

During the MP, participants will remain seated, with their eyes closed and completely still. After receiving guidance on the differences between first-person and third-person perspectives, they will be instructed to use only the first-person perspective, aiming to internally experience each imagined movement.

To support this process, the practice will be guided through structured and standardized verbal instructions, with the goal of maximizing engagement and the effectiveness of motor simulation, as detailed in Table 1. These instructions will be standardized and applied equally to all participants. Their objectives will be: (I) to facilitate vivid and context-rich experiences, encouraging participants to incorporate sensory and environmental details related to the imagined walking scenario; and (II) to maintain participants’ attentional focus on the key gait components presented during the first phase of the training.

**Table 1.**
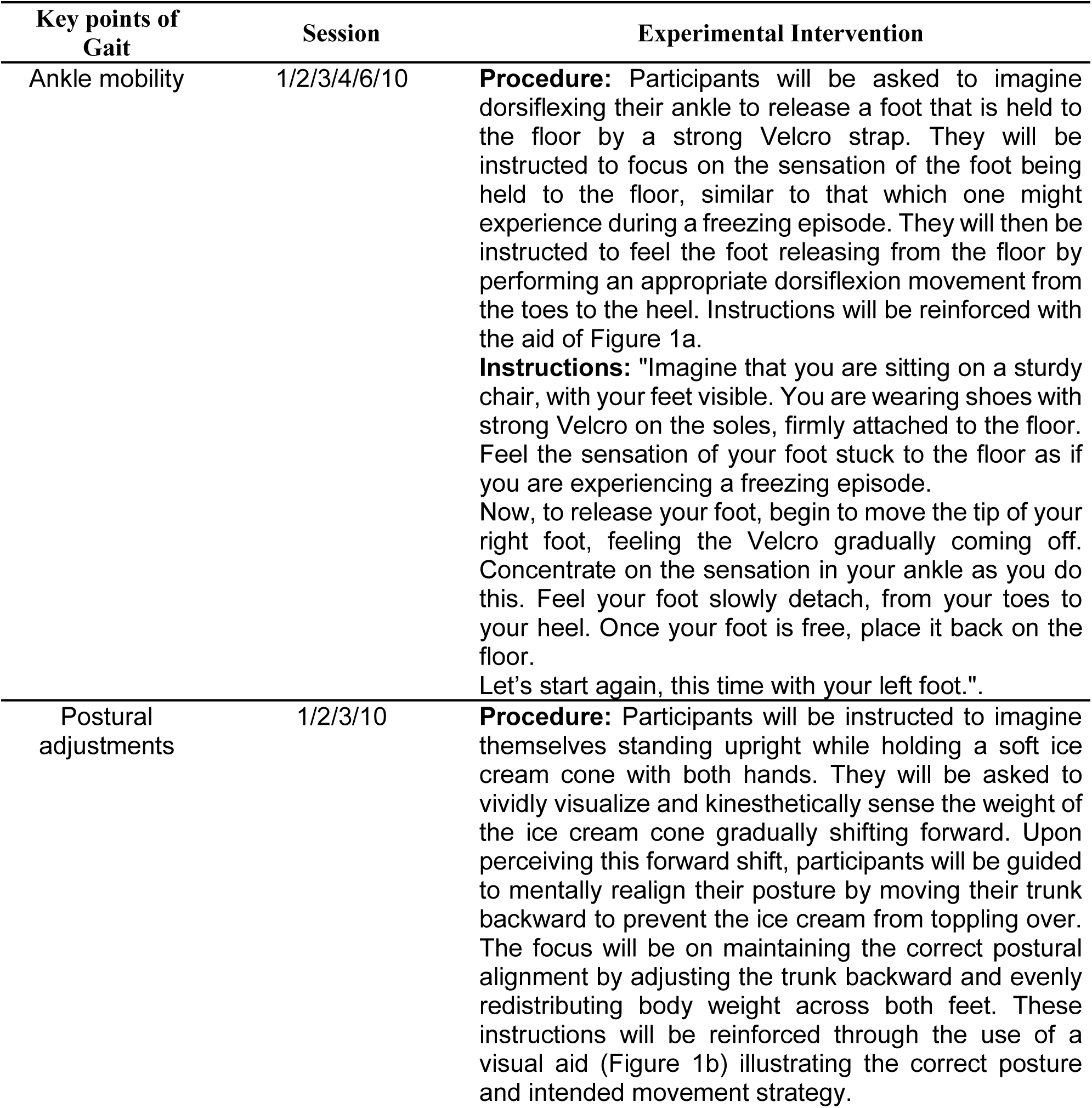
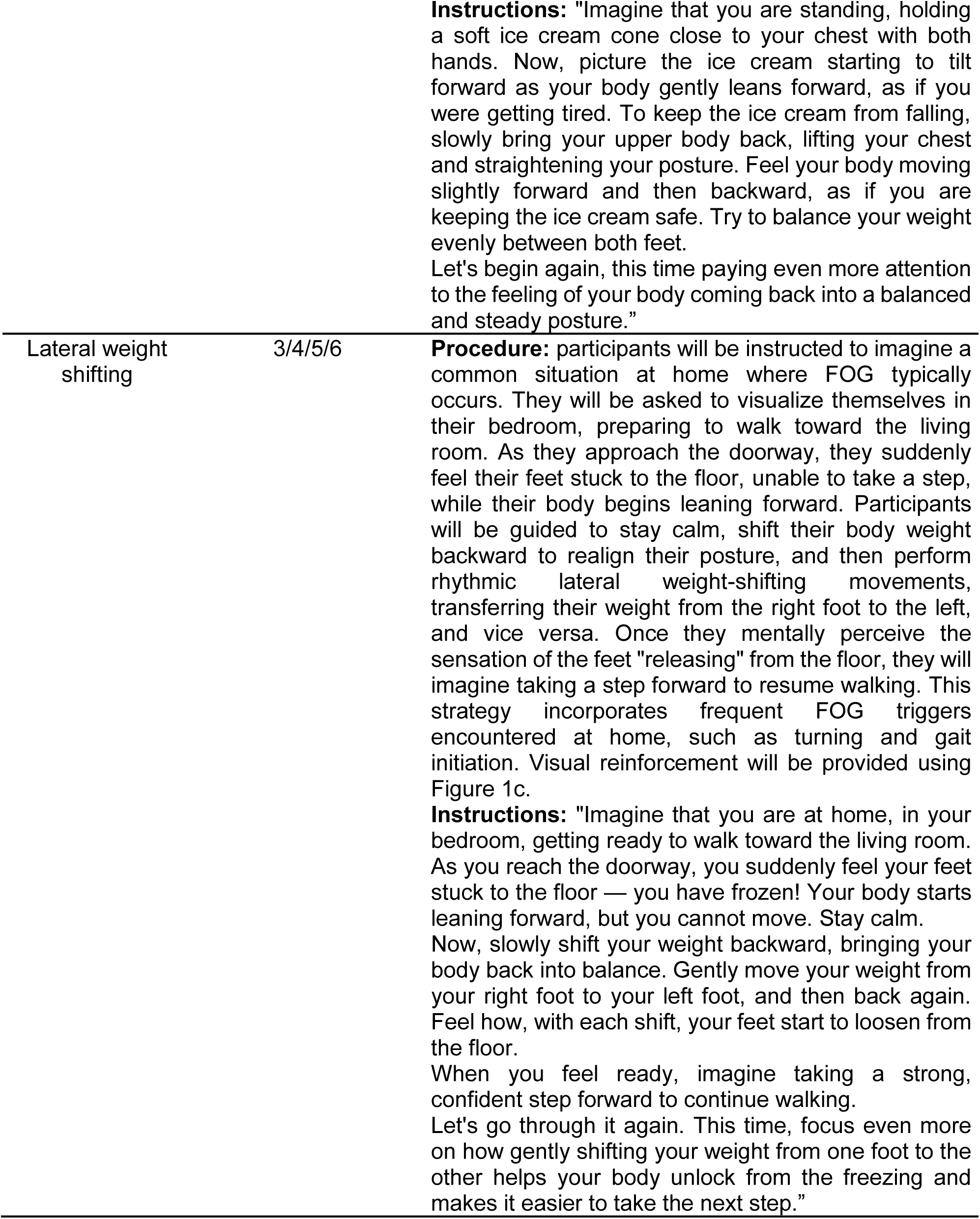
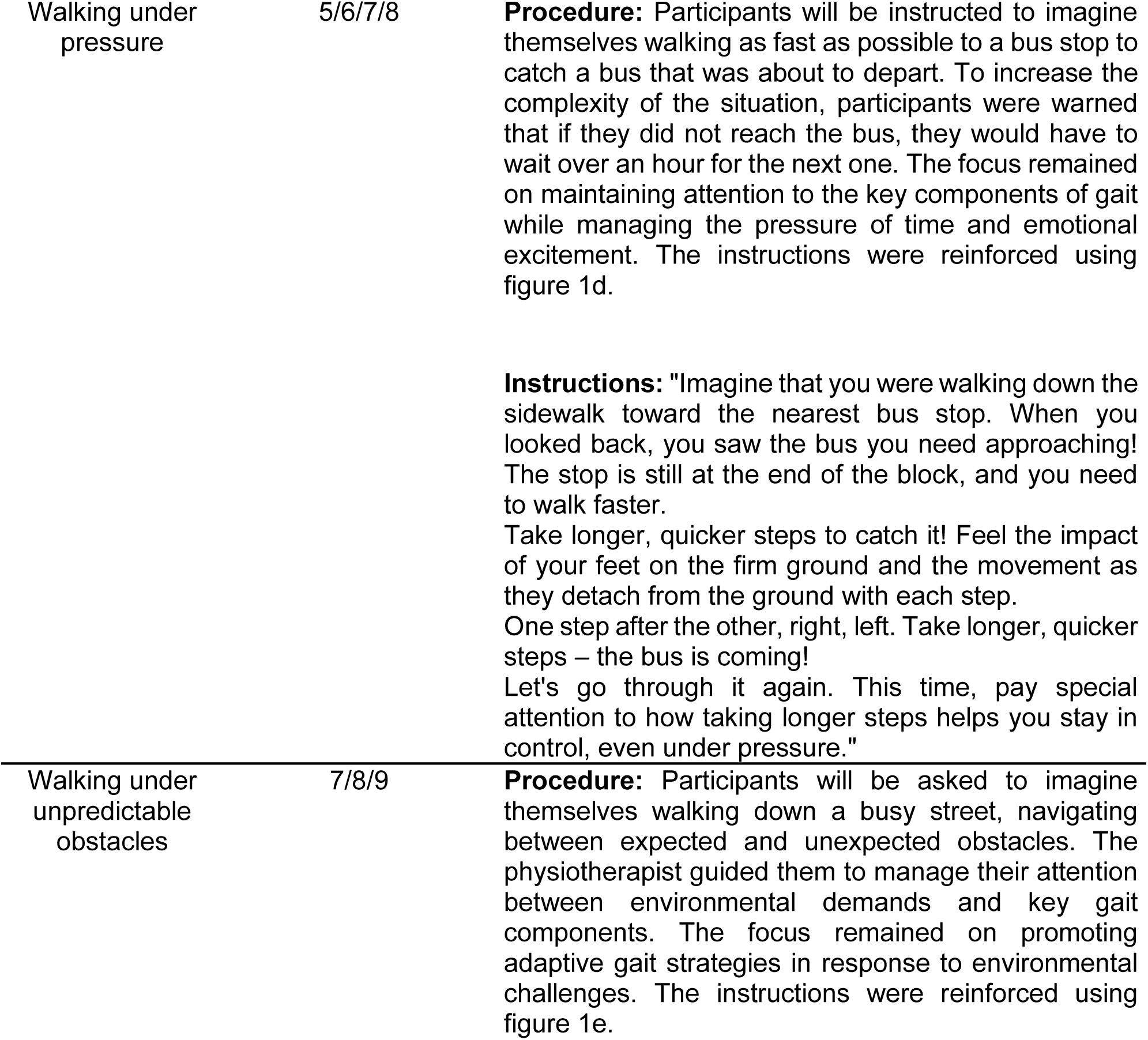
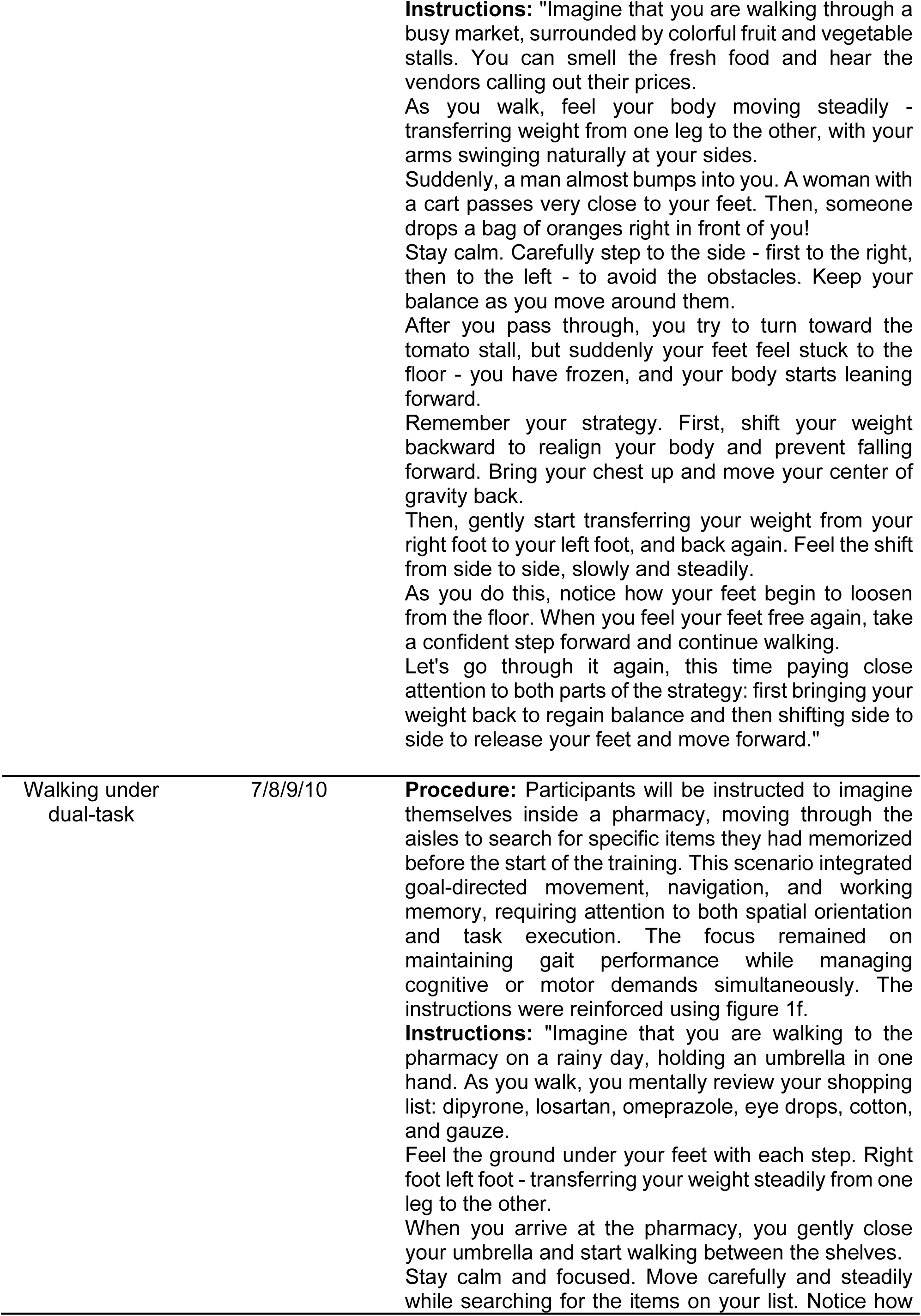
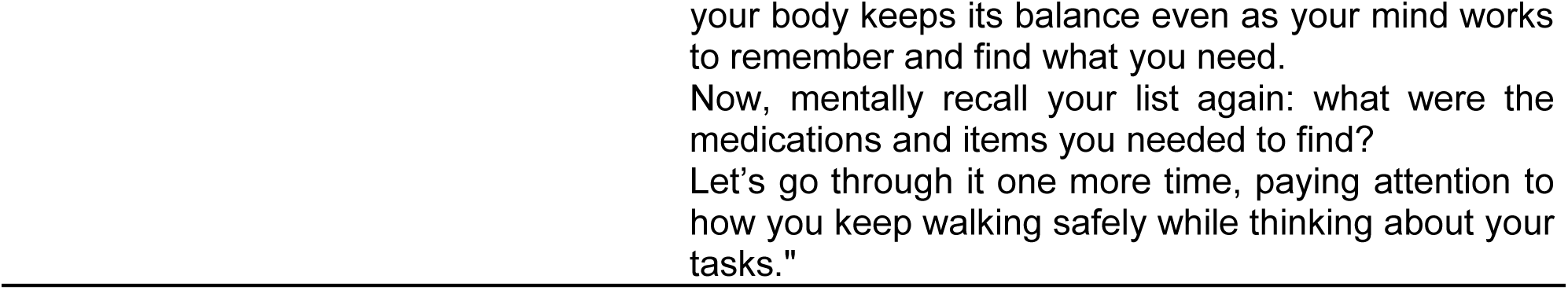
Description of the Experimental Intervention. The table presents each of the key gait components emphasized, the sessions in which they will be addressed, the procedure will be adopted for their execution, and the structured and standardized verbal instructions provided to participants during Mental Practice (MP). FOG = Freezing of Gait.

In addition, illustrations representing each scenario used in MP will be presented before each training block to facilitate imagination (Figure 1).

**Figure 1.**
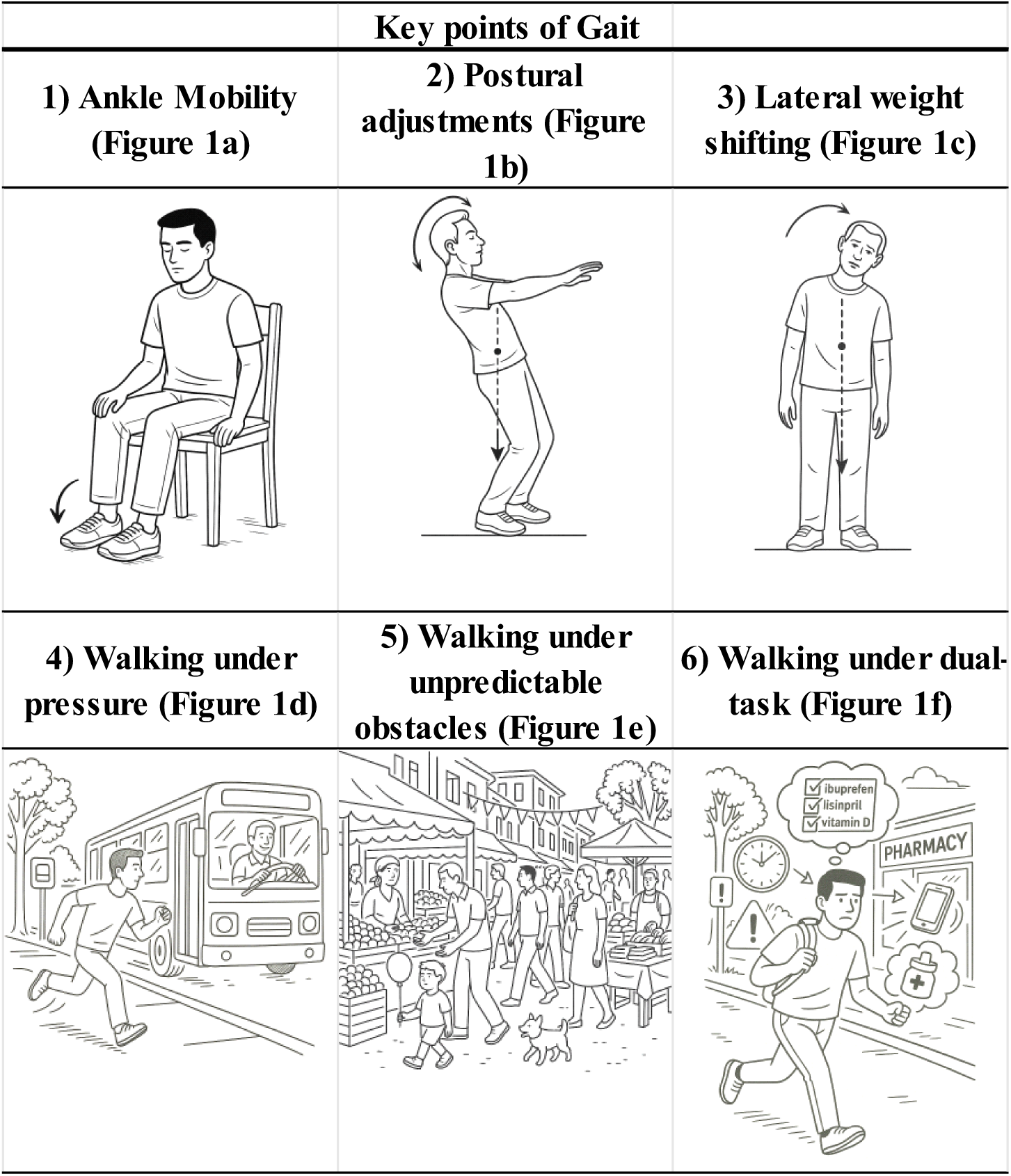
Figures that will be presented to participants to facilitate Mental Practice (MP). Figures that will be used during the experimental intervention to support Mental Practice (MP) by visually reinforcing key gait components and environmental contexts. These visuals will be introduced alongside verbal instructions in each training block.

#### Physical Practice

The PP will consist of performing various exercises selected to simulate functional gait components, with a focus on elements previously addressed in the MP. The activities will involve movements performed in different postures: seated (such as ankle mobility), standing (such as shifting the center of gravity backward and lateral weight transfer), and small displacements (with a maximum distance of 2 meters), as described in Table 2.

**Table 2.**
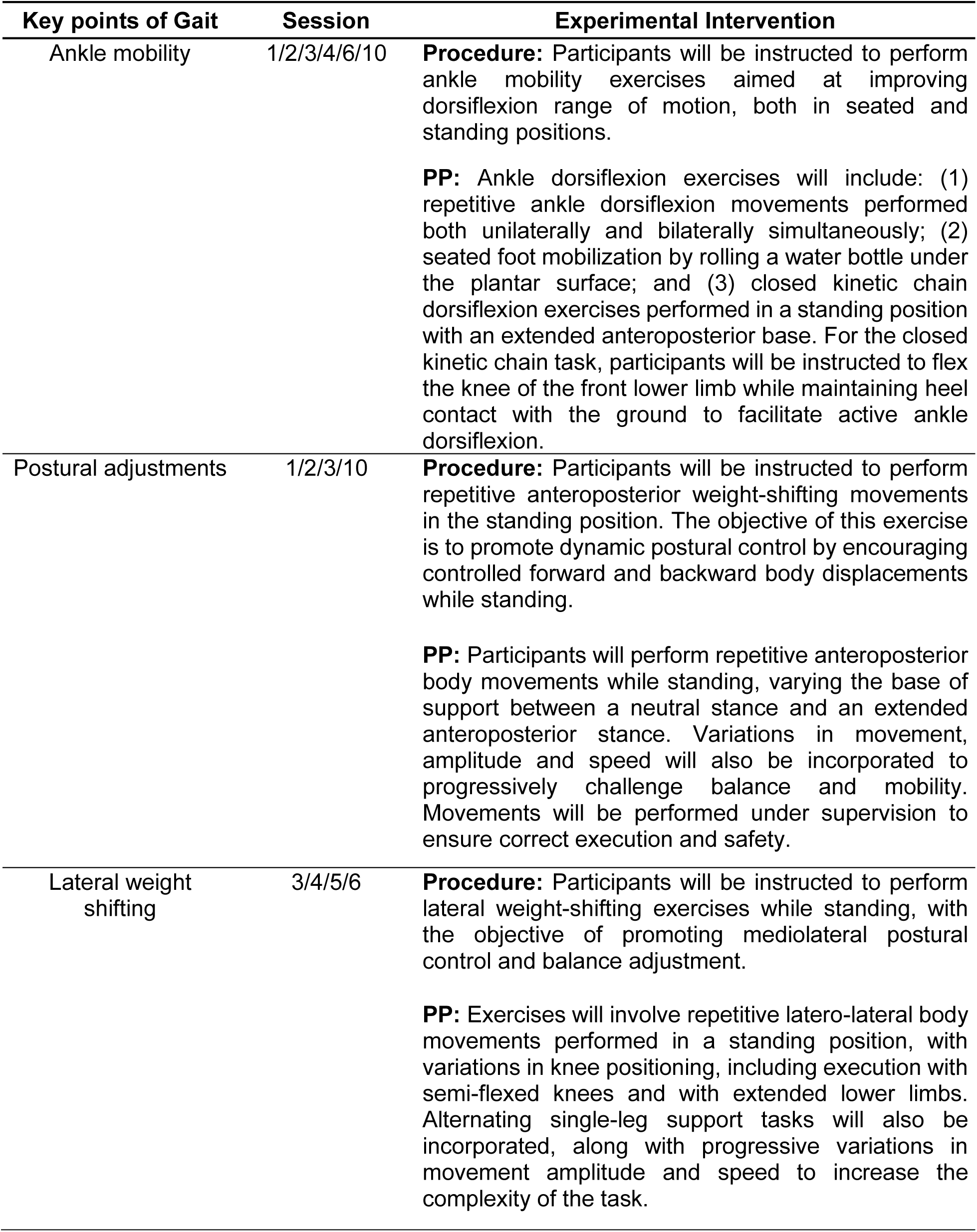
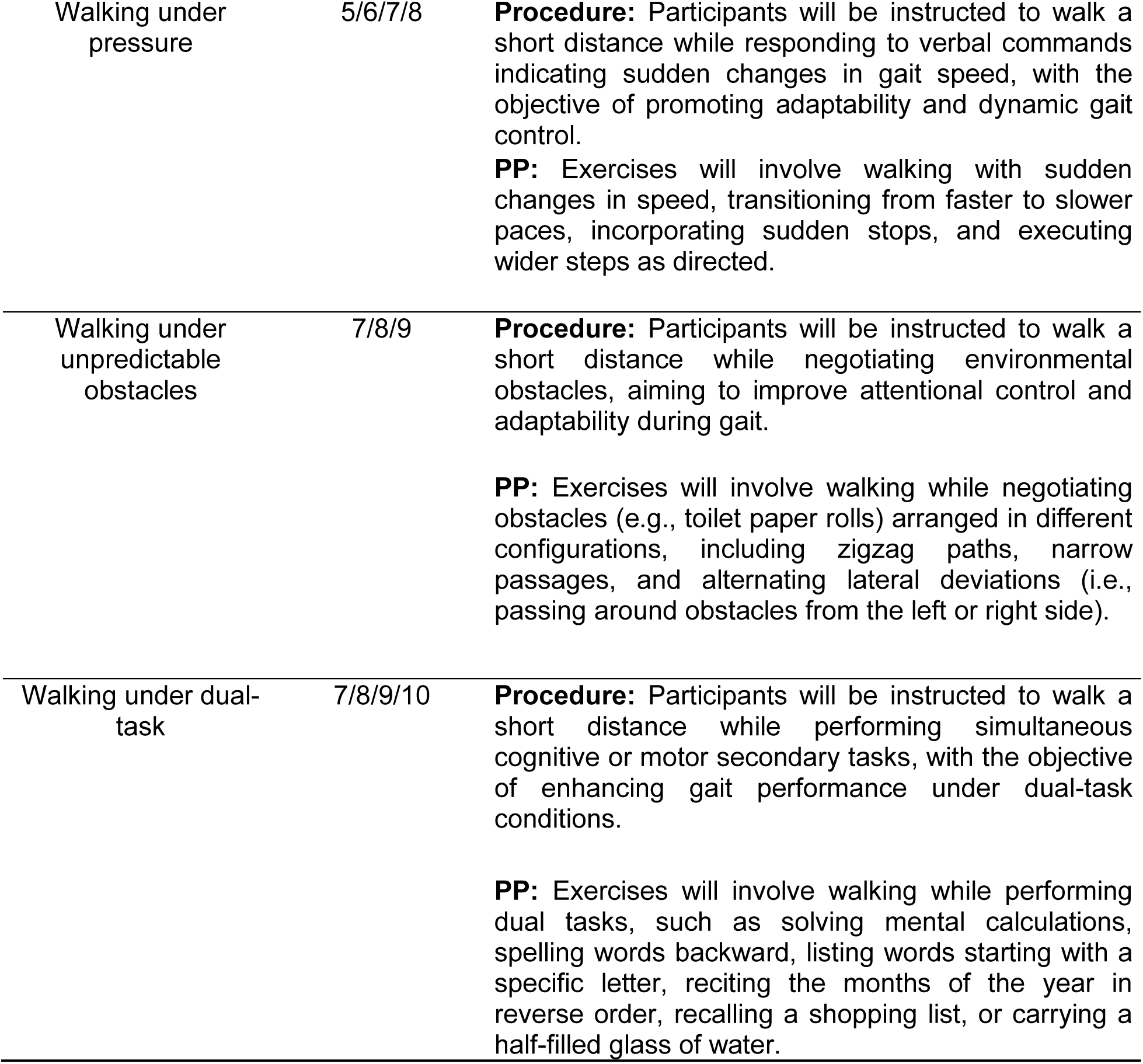
Description of Physical Practice (PP). The table presents each of the key gait components and the sessions in which they will be addressed, the procedure will be adopted for their execution, and the structured and standardized instructions provided to participants during Physical Practice (PP). FOG = Freezing of Gait.

Each exercise will first be demonstrated by the physiotherapist in a clear and detailed manner. Subsequently, the participant’s understanding of the task will be assessed to ensure correct comprehension and readiness for execution. In parallel, all safety conditions will be verified, including confirmation of an obstacle-free environment, the availability of firm support surfaces, appropriate footwear, and, when necessary, the presence of a caregiver or family member to assist. Only after both the participant’s understanding and the environmental safety have been confirmed will the participant be instructed to initiate the exercise.

Throughout the exercise session, the physiotherapist will actively monitor the session via video, having a complete view of the participant’s posture and movements, making real-time adjustments and verbal corrections when needed. The number of repetitions for each exercise will be controlled and standardized according to the protocol. If necessary, rest periods will be offered between exercises. Additionally, the physiotherapist will provide continuous motivational stimuli to encourage adherence and the completion of each task, ensuring the participant’s active engagement in all phases of the PP.

In Table 3, the interaction between the simulated contexts during MP and the exercises addressed in PF is summarized.

**Table 3.**
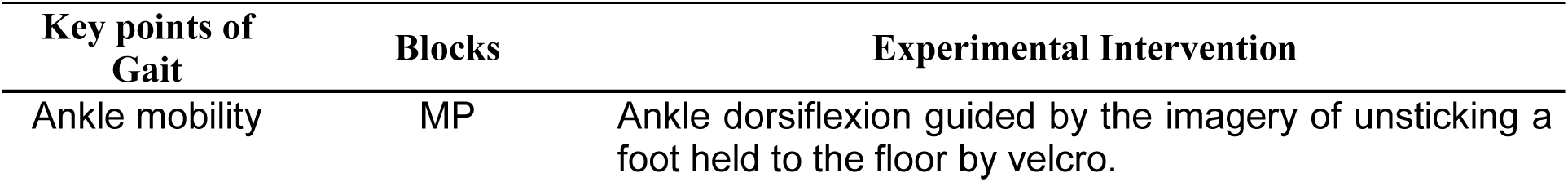
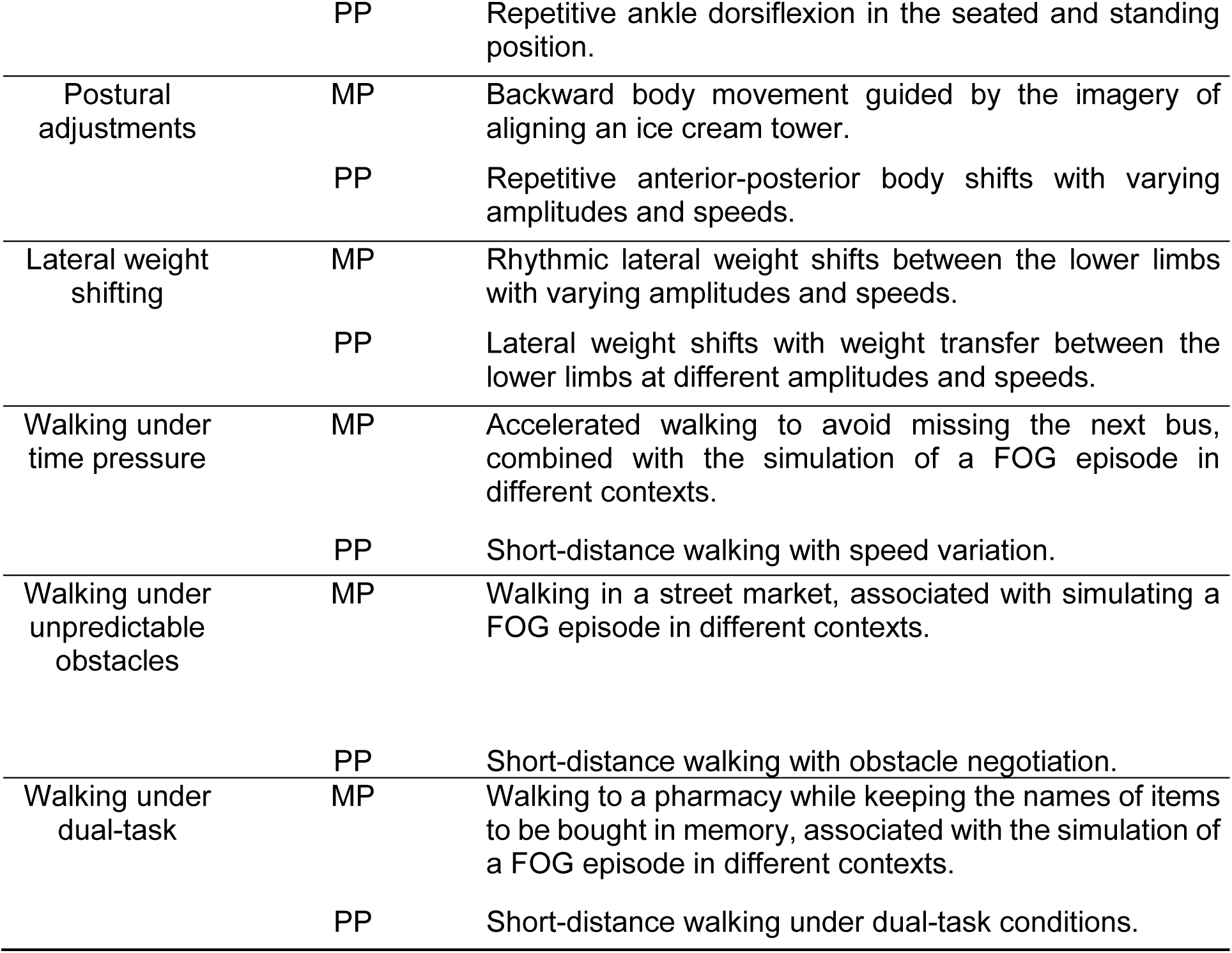
Description of the interaction between Mental Practice (MP) and Physical Practice (PF). The table summarizes the interaction between the contexts that will be worked on in Mental Practice (MP) and the corresponding activities performed in Physical Practice (PF).

| Key points of Gait | Blocks | Experimental Intervention |
| --- | --- | --- |
| Ankle mobility | MP | Ankle dorsiflexion guided by the imagery of unsticking a foot held to the floor by velcro. |
|  | PP | Repetitive ankle dorsiflexion in the seated and standing position. |
| Postural adjustments | MP | Backward body movement guided by the imagery of aligning an ice cream tower. |
|  | PP | Repetitive anterior-posterior body shifts with varying amplitudes and speeds. |
| Lateral weight shifting | MP | Rhythmic lateral weight shifts between the lower limbs with varying amplitudes and speeds. |
|  | PP | Lateral weight shifts with weight transfer between the lower limbs at different amplitudes and speeds. |
| Walking under time pressure | MP | Accelerated walking to avoid missing the next bus, combined with the simulation of a FOG episode in different contexts. |
|  | PP | Short-distance walking with speed variation. |
| Walking under unpredictable obstacles | MP | Walking in a street market, associated with simulating a FOG episode in different contexts. |
|  | PP | Short-distance walking with obstacle negotiation. |
| Walking under dual-task | MP | Walking to a pharmacy while keeping the names of items to be bought in memory, associated with the simulation of a FOG episode in different contexts. |
|  | PP | Short-distance walking under dual-task conditions. |

### Control Intervention

The structure adopted in the CI was designed to ensure equivalence in overall session duration, motor exposure, and organization when compared to the EI. The CI protocol will consist of nine sessions, each structured into four blocks of 10 minutes, totaling 40 minutes per session. Two blocks will involve stretching exercises, while the other two blocks will consist of PP tasks identical to those used in the EI. Thus, the only difference between the interventions will be that the MP blocks implemented in the EI are replaced by stretching exercises in the CI, preserving the total intervention time and motor engagement across groups.

The stretching exercises will target the scapular region, shoulders, elbows, wrists, and fingers, and were specifically chosen because they represent low-intensity physical activities that do not directly influence gait performance or FOG. Participants will be instructed to perform the stretching exercises while seated to maximize safety and minimize postural demands. Each stretch will first be demonstrated clearly and in detail by the physiotherapist. After the demonstration, participant comprehension and safety conditions will be verified, including ensuring the availability of a stable chair and, when necessary, the presence of a caregiver or family member to assist. Only after confirming both understanding and environmental safety will the participant be instructed to initiate the exercises.

Throughout the stretching blocks, the physiotherapist will actively monitor the sessions via video, maintaining a full view of the participant’s posture and movement execution. Real-time verbal feedback and corrective instructions will be provided as needed. The duration of each stretch and the rest intervals between stretches will be standardized according to the study protocol to ensure consistency across sessions and participants.

The PP tasks performed in the CI **will be identical** those of the EI in content, structure, and progression, but without the incorporation of additional cognitive strategies or mental imagery components.

**Table 4.** Description of the Control Intervention (CI). The table presents the structured and pre-defined instructions that will be provided during the Control Intervention (CI) to guide participants stretching exercises. Each exercise will be performed in a seated position with proper posture.

| Key points |  | Control Intervention |
| --- | --- | --- |
| Scapula and Shoulder | 1 | **Starting Position:** Seated with the back supported by a chair, head aligned with the shoulders, elbows extended, hands resting on the thighs above the knees, and feet flat on the floor.<br>**Execution:** While maintaining an upright posture, perform a shoulder depression movement toward the pelvis, sliding the hands forward along the thighs without lifting them. |
| Scapula and Shoulder | 2 | **Starting Position:** Seated with the back supported by a chair, head aligned with the shoulders, and feet flat on the floor.<br>**Execution:** Cross the upper limbs so that each hand rests on the posterolateral region of the opposite shoulder. Then, flex the trunk slightly forward, bringing the elbows forward in front of the body. |
| Shoulder | 3 | **Starting Position:** Seated with the back supported by a chair, head aligned with the shoulders, and feet flat on the floor.<br>**Execution:** While maintaining an upright posture, flex the shoulders and elbows to position the hands behind the neck with fingers interlaced. Then, gently move the elbows backward, projecting the chest forward. |
| Shoulder | 4 | **Starting Position:** Seated with the back supported by a chair, head aligned with the shoulders, and feet flat on the floor.<br>**Execution:** While maintaining an upright posture, flex the shoulder and elbow to bring the palm of the hand toward the upper region of the ipsilateral scapula. Using the opposite hand, apply gentle backward pressure to deepen the stretch. |
| Elbow, wrist, and fingers | 5 | **Starting Position:** Seated with the back supported by a chair, head aligned with the shoulders, elbows extended, hands placed on a stable surface beside the body with fingers pointing backward, and feet flat on the floor.<br>**Execution:** Lean the torso backward while maintaining hand contact with the support surface, promoting shoulder extension. |
| Elbow, wrist, and fingers | 6 | **Starting Position:** Seated with the back supported by a chair, head aligned with the shoulders, hands placed on a stable surface in front of the body, and feet flat on the floor.<br>**Execution:** Perform radial deviation of the wrist without lifting the hand from the support surface. |
| Elbow, wrist, and fingers | 7 | **Starting Position:** Seated with the back supported by a chair, head aligned with the shoulders, hands placed on a stable surface in front of the body, and feet flat on the floor.<br>**Execution:** Perform ulnar deviation of the wrist without lifting the hand from the support surface. |
| Elbow, wrist, and fingers | 8 | **Starting Position:** Seated with the back supported by a chair, head aligned with the shoulders, and feet flat on the floor.<br>**Execution:** While maintaining an upright posture, elevate the arms to 90° of shoulder flexion with elbows extended, so that the palms face the torso. Using the opposite hand, apply gentle backward pressure to the dorsal aspect of the fingers, bringing them toward the forearm. |
| Elbow, wrist, and fingers | 9 | **Starting Position:** Seated with the back supported by a chair, head aligned with the shoulders, and feet flat on the floor.<br>**Execution:** While maintaining an upright posture, flex both elbows to 90° at shoulder height, bringing the palms together with fingers pointing upward in front of the body. Apply gentle pressure between the palms while moving the wrists downward toward the floor. |
| Elbow, wrist, and fingers | 10 | **Starting Position:** Seated with the back supported by a chair, head aligned with the shoulders, and feet flat on the floor.<br>**Execution:** While maintaining an upright posture, perform internal shoulder rotation with 90° of elbow flexion (positioning the forearm in front of the torso) and flex the wrist so that the fingers point downward. Using the opposite hand, apply gentle backward pressure to the dorsal surface, bringing the fingers toward the forearm. |

### Criteria for discontinuing or modifying allocated interventions {11b}

The intervention will be discontinued upon the participant’s request and/or abandonment of the prescribed sessions and evaluations, situations where there is a worsening of the progressive disease leading to an inability to maintain an upright posture and walk unassisted, as well as in cases of concomitant participation in other new interventions or treatments that may modify the analyzed outcomes.

### Strategies to improve adherence to interventions {11c}

To promote session adherence, participants will receive a reminder of the scheduled date and time one day prior. The protocol is not anticipated to cause significant adverse effects, with the exception of potential physical and/or mental fatigue, that is expected to resolve quickly after resting in a seated position. Any adverse events or complaints arising during the training related to the protocol will be tracked. Additionally, serious adverse events, such as fractures, hospitalizations, or surgeries that prevent continuation of training, will be closely monitored.

### Relevant concomitant care and interventions that are permitted or prohibited during the trial {11d}

To ensure continuity of training, the participant will not be able to participate in other new interventions or treatments that may modify the analyzed outcomes.

### Provisions for post-trial care {30}

No significant adverse effects are expected as a result of the established protocol, only a possible sensation of physical and/or mental fatigue that is expected to resolve quickly after resting in a seated position.

### Outcomes {12}

The two groups will be evaluated at three different times: an assessment before intervention (BI), after intervention (AI) and 30 days after the intervention as a follow-up (FU). Demographic data (date of birth, age, gender, education and profession) will be collected during the first assessment.

#### Primary outcomes

1. Rapid Turns Test: a provocative freezing test. The individuals will be instructed to spin around their own axis repeatedly, in both directions, at high speed [40]. This is a reliable, valid, and sensitive measure for investigating the presence of FOG, as spinning remains the most reliable trigger for inducing the symptom [41].

Before the test was conducted, the evaluator will ensure that the environment is appropriate, instructing the participant - and, when available, their caregiver or family member - to organize the space in such a way that complete turns around their own axis, in both directions, can be performed without obstacles or rugs, and with sufficient lighting. The participant will be instructed to position themselves close to a stable support (such as a wall, countertop, or stable chair) to be used in case of imbalance.

Next, the evaluator will explain in detail the purpose of the test, the execution criteria, and will demonstrate the task to the participant. The participant will be instructed that, upon hearing the verbal cue “Now,” they should begin turning on their own axis, alternating between the right and left side with each full turn, as quickly as possible and continuously, until they are asked to stop. It will be reinforced that, in case of imbalance, the available support should be used immediately, and that difficulties or freezing episodes will be expected and will not be considered errors. The evaluator will ensure that all questions are answered and that the participant feels comfortable and safe to perform the test.

The test will only begin after explicit confirmation that the participant has understood all instructions and feels safe to perform it. Throughout the evaluation, the physiotherapist will have a clear view of the participant’s body and will actively supervise the task, providing continuous verbal support. The performance will be recorded for later analysis.

1. II) Percentage of time spent with FOG during the task (%FOG) - A visual scoring of FOG episodes will be performed from video recordings to measure the actual severity of freezing, calculated using the formula: (total duration of FOG during the test * 100 / total duration of the test).

#### Secondary outcomes

1. New Freezing of Gait Questionnaire (NFOG-Q): the participants will sit while completing the self-reported questionnaire that assesses the clinical aspects of FOG and the impact on quality of life. The total scores range from 0 to 28, with higher scores indicating greater severity impact of FOG [37].
2. Movement Disorders Society - Unified Parkinson’s Disease Rating Scale (MDS-UPDRS): It is a widely recognized scale used to evaluate and track the progression of Parkinson’s Disease (PD). The scale includes 42 items, categorized into four sections: Part I (non-motor aspects of daily living), Part II (motor aspects of daily living), Part III (motor evaluation), and Part IV (motor complications). The participants will sit while completing Part II (motor aspects of daily living), consisting of 13 questions. The score ranges from 0 to 4 for each item, with the maximum value indicating greater impairment in daily living activities [42].
3. Parkinson Disease Questionnaire - 39 (PDQ-39): the participants will sit while completing a questionnaire consisting of 39 questions distributed across eight domains (mobility - ten items; activities of daily living - six items; emotional well being - six items; social support - three items; physical discomfort - three items; stigma - four items; and communication - three items). Each item can be answered with one of five predetermined responses (never, rarely, sometimes, frequently, and always). The score for each item ranges from 0 to 4, with the total score ranging from 0 to 100, where the lowest score reflects better quality of life [43].
4. Telephone Montreal Cognitive Assessment (T-MoCA): The instrument evaluates six cognitive domains, including naming, memory, attention, language, abstraction, and orientation. The participants will sit while completing the cognitive test that does not require the use of paper, pencil or visual stimuli. The maximum score is 22 points [38].

#### Participant timeline {13}

**Figure 2.**
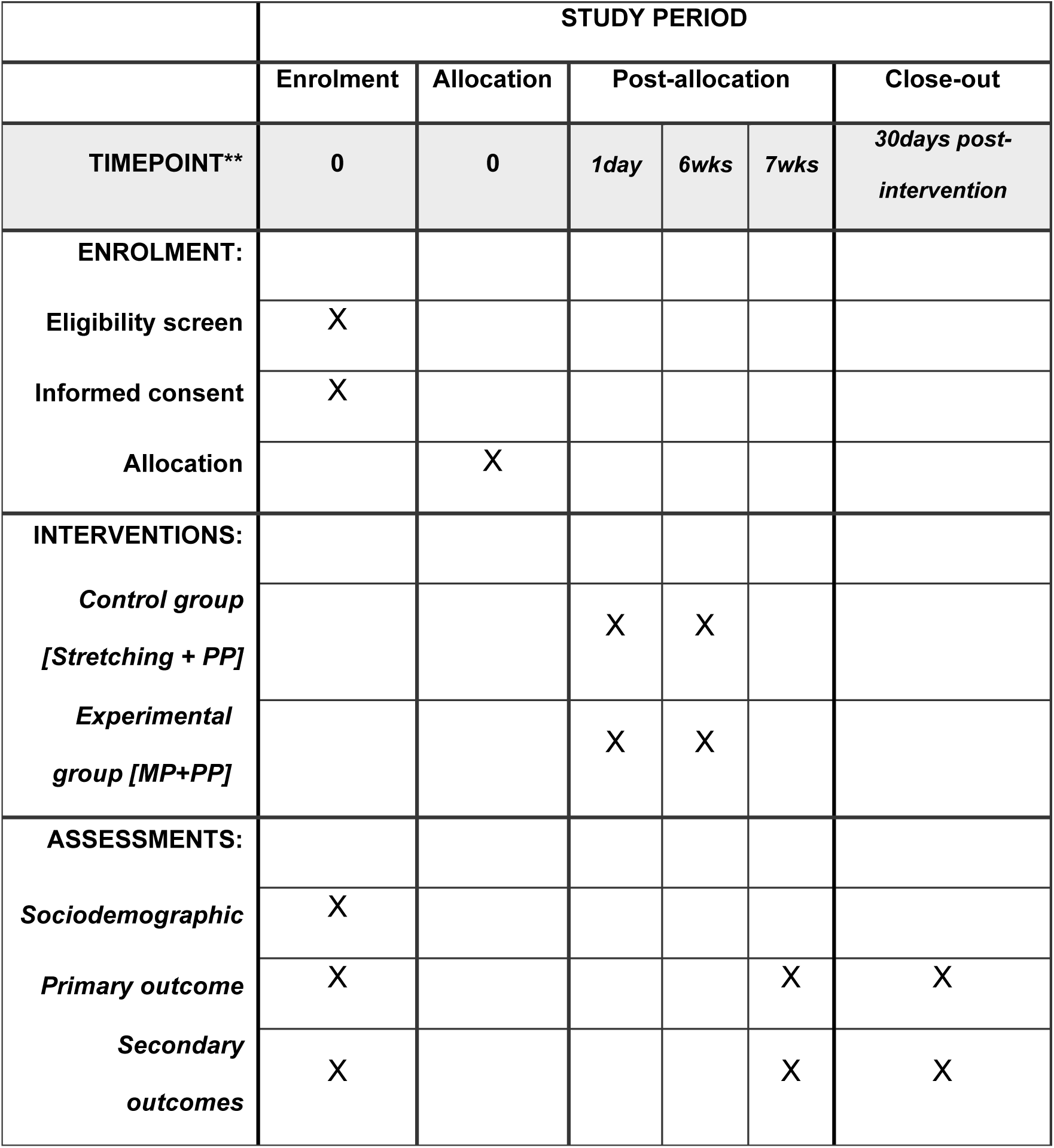
The schedule of enrollment, interventions and assessments demonstrated in the Standard Protocol Items: Recommendations for Interventional Trials (SPIRIT) figure. This figure presents the timeline for participant enrollment, the sequence of interventions, and the assessment points throughout the study, following the SPIRIT guidelines for clinical trials. It outlines the key phases of the trial, from initial screening to follow-up evaluations, ensuring a clear overview of the study protocol and participant progression.

**Figure 3.**
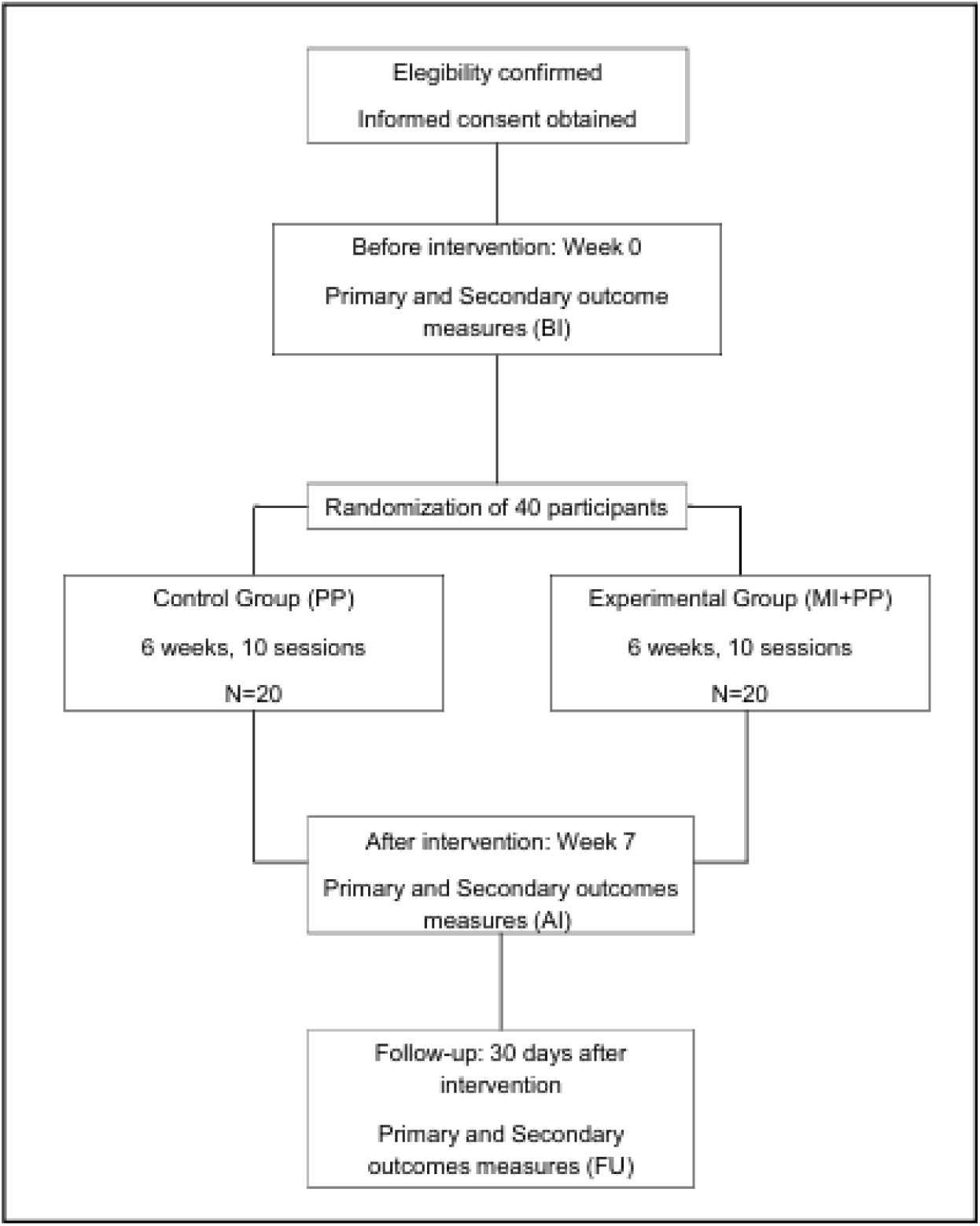
The schematic study design. This figure illustrates the overall structure of the study, showing the various stages and processes involved, including participant recruitment, intervention phases, and assessment points.

#### Sample size {14}

The sample size calculation was based on the secondary outcome, NFGO-Q. Considering an effect size (ES) of 0.50 based on a previous study that reported a significant interaction effect for NFOG-Q scores [44], a significance level of 0.05, a desired power of 0.80, and a correlation of 0.5 between repeated measures, the power analysis determined that 14 individuals per group are required. Considering possible sample losses, the final sample will consist of 50 individuals.

### Recruitment {15}

Participants are being recruited consecutively from the contacts of the AMPARO network (www.amparo.numec.prp.usp.br) using a non-probability sampling method. Recruitment began in January 2025, and it’s currently ongoing.

### Assignment of interventions: allocation

#### Sequence generation, concealment mechanism and implementation {16a/16b/16c}

Participants will be randomly assigned to the experimental or control group using a computer-generated simple randomization sequence, with an allocation ratio of 1:1, generated by an independent researcher not involved in recruitment, training or assessment.

Only the researcher responsible for conducting the training will know the participant’s allocation. The outcome assessor, responsible for the initial assessments and reassessments, will remain unaware of the allocation throughout the data collection process, staying blinded to the type of intervention offered to each participant during the entire study. Additionally, participants will be instructed to not discuss details of the training with the evaluator.

#### Who will be blinded {17a}

The outcome assessor, responsible for the assessments and reassessments, will not be informed by the researcher responsible of participant’s allocation during data collection, and the participants will be instructed to not discuss details of the training with the evaluator.

#### Procedure for unblinding {17b}

The outcome assessor will only become aware of the allocation of participants after the entire data collection stage has been completed, and then the final data analysis stage has begun.

### Data collection and management

#### Plans for assessment and collection of outcomes {18a}

All assessments will be conducted remotely via encrypted video call by a single physical therapist with clinical specialization in movement disorders. The outcome assessor will be blinded to group allocation and will have no involvement in delivering the interventions. Prior to the start of data collection, the assessor will undergo structured training for the administration of all selected assessment tools, as well as for the standardization of procedures, in order to ensure consistency and minimize measurement bias.

Assessments will be scheduled in advance and conducted at the same time of day for each participant, within their self-reported dopaminergic medication ON period. This strategy will aim to reduce the influence of medication-related motor fluctuations on assessment performance. Participants will be instructed to ensure a quiet and well-lit environment with proper camera positioning to optimize visibility and measurement accuracy during remote assessments.

#### Plans to promote participant retention and complete follow-up {18b}

There is no financial or other type of reward for participating in the study. To ensure complete follow-up, participants will be contacted frequently to remind them of the dates and times scheduled for assessments and sessions.

#### Data management {19}

Only the researchers involved will have access to the data collected and information from the participants. In order to guarantee the security of personal information, each participant will receive an alphanumeric code.

#### Confidentiality {27}

The outcomes assessor will have contact with the participants and their personal information only during the assessments, and at the time the participant will be instructed to not mention details of the procedure that they submitted during the intervention sessions.

Regarding the data obtained during the collection, they will not be disclosed under any circumstances except within the scientific scope.

#### Plans for collection, laboratory evaluation and storage of biological specimens for genetic or molecular analysis in this trial/future use {33}

Not applicable.

## Statistical methods

### Statistical methods for primary and secondary outcomes {20a}

In case of missing data in the AI and FU assessments of participants who may be lost during the study, data imputation will be performed using the mean of the respective groups. This method will be adopted because it provides the most consistent results with the least variability, ensuring the preservation of the data structure and maintaining the representativeness of each group’s characteristics.

Group comparisons at baseline (BI) will be conducted using parametric and non-parametric tests, depending on the nature and distribution of the variables. For variables that present a normal distribution (verified using the Shapiro-Wilk test), the independent Student’s t-test will be applied. For all other variables that do not present a normal distribution, the non-parametric Mann-Whitney U test will be used.

To evaluate the effects of the intervention across time points, appropriate statistical tests will be selected based on the distributional characteristics of the data. For normally distributed (parametric) data, a repeated-measures ANOVA will be conducted, followed by Tukey’s post-hoc test for multiple comparisons. For non-normally distributed (non-parametric) data, a permutation-based ANOVA with 10,000 permutations will be applied, followed by a Bonferroni-corrected post-hoc test. The permutation-based approach was chosen for its robustness and its ability to handle potential violations of standard parametric assumptions, including normality and homogeneity of variance.

A linear trend analysis will also be conducted to assess the pattern of change in %FOG across the three assessments (BI, AI, and FU) within each group. For this analysis, linear regression coefficients, coefficients of determination (R²), and p-values will be calculated separately for each group. The significance level adopted for all analyses will be 5% (p < 0.05).

All statistical analyses will be conducted using Python software (version 3.11), with a significance level set at 95%.

#### Interim analyses {21b}

Not applicable.

#### Methods for additional analyses (e.g. subgroup analyses) {20b}

Not applicable.

### Methods in analysis to handle protocol non-adherence and any statistical methods to handle missing data {20c}

Not applicable.

#### Plans to give access to the full protocol, participant level-data and statistical code {31c}

Not applicable.

### Oversight and monitoring

#### Composition of the coordinating centre and trial steering committee {5d}

Not applicable.

#### Composition of the data monitoring committee, its role and reporting structure {21a}

Not applicable.

#### Adverse event reporting and harms {22}

Adverse events or complaints that occurred during training sessions that are related to the protocol will be monitored. Serious adverse events, such as fractures, hospitalizations, or surgeries that prevent continuation of training, will be closely monitored.

#### Frequency and plans for auditing trial conduct {23}

Not applicable.

#### Plans for communicating important protocol amendments to relevant parties (e.g.trial participants, ethical committees) {25}

If there is a need for important protocol modification, the relevant parties will be communicated as far in advance as possible.

### Dissemination plans {31a}

The results will be published in journals after completion of the study. A simplified summary will be made available to participants and their families, in addition to being shared through the AMPARO platform and its respective social media channels.

## Discussion

FOG is a highly disabling phenomenon in PD strongly associated with the progressive deterioration of automatic gait control mechanisms [6,7,8]. As the functional connectivity of the basal ganglia-thalamocortical circuits declines, people with PD progressively rely on attentional and executive control to compensate for reduced locomotor automaticity [7,8]. Neuroimaging studies have shown that people with PD and FOG show increased activation in prefrontal and parietal areas during gait and motor imagery tasks [10,14], reflecting a compensatory recruitment of cortical networks to sustain gait initiation and execution.

The novel proposed intervention combining MP based on DNI and PP is specifically designed to improve these compensatory mechanisms. Structured MP engages frontoparietal circuits involved in motor planning, postural control, and visuospatial processing [17,18,19]. Particularly, kinesthetic, first-person perspective imagery - a core element of DNI - has been shown to activate cortical networks similar to those observed during actual movement execution [17,20,27]. Repeated engagement of such imagery tasks may promote neuroplastic changes in executive-attentional networks reducing the impacts of insufficient automatic motor control [10,11,12,14].

Moreover, by integrating goal-directed control, obstacle negotiation, dual-task conditions, and adaptive postural strategies into the practice, the intervention trains participants to proactively manage environmental demands and internal motor planning [9,13,21]. This approach directly targets deficits commonly implicated in FOG, such as impaired set-shifting, reduced internal cue generation, and increased susceptibility to environmental triggers [8,9]. This novel intervention may enhance gait performance and decrease FOG severity by improving gait attentional control.

In addition, another advantage of the proposed intervention is its remote delivery format, which minimizes critical barriers often faced by people with PD, such as reduced mobility, limited access to specialized rehabilitation services, and transportation challenges [32,34]. By reducing logistical and geographical obstacles, remotely delivered interventions have been shown to enhance accessibility, promote higher adherence to rehabilitation programs, and reduce overall healthcare costs [32,34]. This approach is particularly valuable for individuals living in remote areas or those with significant motor impairments, supporting a more inclusive and sustainable model of care for people with PD. Besides, the novel intervention does not demand specialized equipment, wearable devices, or high-cost technological resources [32,33].

Furthermore, by emphasizing internal cognitive-motor strategies rather than external cue dependence, the intervention promotes long-term self-management skills, empowering patients to actively apply compensatory strategies in their daily living activities [8,32,33]. This is consistent with current rehabilitation guidelines that emphasize adaptability, autonomy, and resilience as important goals in the management of chronic neurological conditions [32,33].

Thus, the intervention proposed here not only addresses the pathophysiological mechanisms underlying FOG but also offers a scalable, accessible, cost-effective model for cognitive-motor rehabilitation in PD. If successful, it has the potential to significantly impact clinical practice by expanding access to effective gait rehabilitation strategies and improving the quality of life for people with PD worldwide.

### Strengths and Limitations

This trial presents several methodological strengths. It addresses a critical gap in the literature by targeting FOG in PD through a novel, structured, remote intervention combining MP based on DNI and PP. The protocol integrates goal-directed, cognitively demanding gait tasks that simulate real-world challenges, enhancing ecological validity. It also uses a remote, low-cost delivery model that increases accessibility, scalability, and feasibility for diverse healthcare settings.

Limitations include potential challenges related to adherence monitoring in a fully remote intervention format and the reliance on self-reported measures for some secondary outcomes, which may introduce bias. Additionally, the generalizability of findings may be limited to individuals with mild to moderate PD who are able of participating in online sessions. These considerations will be addressed through structured adherence tracking, blinded outcome assessments, and comprehensive reporting of participant characteristics.

## Conclusion

This study protocol proposes a novel randomized controlled trial designed to evaluate the effects of a novel remote intervention combining MP based on DNI and PP on FOG in people with PD. By targeting attentional control mechanisms and promoting cognitive-motor adaptability, the intervention has the potential to compensate for impaired gait automaticity, improving functional mobility and quality of life. Additionally, its low-cost, scalable, and accessible format offers a promising model for expanding rehabilitation services for PD. The results of this study will contribute with new evidence that could guide the development of innovative gait rehabilitation strategies.

## Trial Status

The study is currently ongoing, with recruitment initiated in January 2025. Approximate recruitment completion date is August 2027.

## Supporting information

PRS Draft Review

## Data Availability

All data produced in the present work are contained in the manuscript.

## Abbreviations

PD: Parkinson’s disease
FOG: Freezing of Gait
AOT: Action Observation Training
MP: Mental Practice
PP: Physical Practice
fMRI: Functional Magnetic Resonance Imaging
TUG: Time UP and Go
DNI: Dynamic Neuro-Cognitive Imagery
CI: Control Intervention
SPIRIT: Standard Protocol Items: Recommendations for Interventional Trials
LEED: Levodopa Equivalent Daily Dose
NFOG-Q: New Freezing of Gait Questionnaire
T-MoCA: Telephone Montreal Cognitive Assessment
KVIQ-20: Kinesthetic and Visual Imagery Questionnaire - 20
EI: Experimental Intervention
BI: Before Intervention
AI: After Intervention
FU: Follow-up
%FOG: Percentage of time spent with FOG during the task
MDS-UPDRS: Movement Disorders Society - Unified Parkinson’s Disease Rating Scale
PDQ-39: Parkinson Disease Questionnaire - 39
ES: Effect Size
REDE AMPARO: Neuromat Support Network for Friends and Individuals with PD
EG: Experimental Group
CG: Control Group

## Acknowledgements

This study was funded by the São Paulo Research Foundation (FAPESP) through the Research, Innovation and Dissemination Center for Neuromathematics (CEPID NeuroMat, grant number 2013/07699-0).

The authors sincerely thank all participants for their collaboration and commitment, as well as the technical and administrative teams who provided essential support in carrying out the research activities. We also acknowledge the institutions involved for their continuous support in the development of this work.

## Declarations

### Author’s contributions {31b}

PRS: study concept, study design, recruitment and screening of participants, preparation and drafting of study protocol, and drafting of manuscript; KYTH: study concept, study design, recruitment and screening of participants, development of intervention; MRP: study concept, study design, development of intervention; ARS: study concept, study design, development of intervention; MEEP: study concept, study design, preparation and drafting of study protocol, drafting and review of manuscript. All authors read and approved the final manuscript.

### Funding {4}

This study is funded by the São Paulo Research Foundation (FAPESP) through the Research, Innovation and Dissemination Center for Neuromathematics (CEPID NeuroMat, grant number 2013/07699-0).

### Availability of data and materials {29}

Not applicable.

### Ethics approval and consent to participate {24}

This project was approved by the Research Ethics Committee of the Faculty of Medicine of the University of Sao Paulo, under protocol number 7.253.458. The study has been registered on ClinicalTrials.gov and is currently awaiting the assignment of the NCT number following review. Attached is the official submission receipt (PRS Draft Receipt). All participants, before the start of the intervention, will be informed about the research and will sign an informed consent form sent online, only then will they participate in the study.

### Consent for publication {32}

Prior to the beginning of the session, all participants will be informed about the research and will sign a free and informed consent form, authorizing the analysis and publication of the data obtained from their participation. Only then will they participate in the study.

### Competing interests {28}

The authors declare that they have no competing interests.

## Notes

### Competing Interest Statement

The authors have declared no competing interest.

### Clinical Trial

The study has been registered on ClinicalTrials.gov and is currently awaiting the assignment of the NCT number following review. Attached is the official submission receipt (PRS Draft Receipt).

### Funding Statement

This study is funded by the Sao Paulo Research Foundation (FAPESP) through the Research, Innovation and Dissemination Center for Neuromathematics (CEPID NeuroMat, grant number 2013/07699-0). FAPESP had no role in the study design, data collection, management, analysis, or interpretation of the data; writing of the protocol; or the decision to submit the results for publication. All scientific and operational decisions were made independently by the authors of the protocol.

### Author Declarations

The Research Ethics Committee of the Faculty of Medicine of the University of Sao Paulo gave ethical approval for this work (number 7.253.458).

