## Supplementary material for "Effects of a Remote Mental and Physical Practice Intervention on Freezing of Gait in People with Parkinson’s Disease: a Single-Blind Randomized Controlled Trial Protocol": PRS Draft Review

**ClinicalTrials.gov PRS DRAFT Receipt (Working Version)**
Last Update: 04/24/2025 15:03

**ClinicalTrials.gov ID: [Not yet assigned]**

### Study Identification

Unique Protocol ID: PM-DP NeuroMat

Brief Title: Remote Mental Practice for Freezing of Gait in Parkinson's Disease: A Randomized Controlled Trial

Official Title: Effects of a Remote Mental and Physical Practice Intervention on Freezing of Gait in People With Parkinson's Disease: a Randomized Controlled Trial

Secondary IDs:

### Study Status

Record Verification: April 2025

Overall Status: Recruiting

Study Start: January 25, 2025 [Actual]

Primary Completion: August 2025 [Anticipated]

Study Completion: January 2028 [Anticipated]

### Sponsor/Collaborators

Sponsor: University of Sao Paulo General Hospital

Responsible Party: Sponsor

Collaborators:

### Oversight

U.S. FDA-regulated Drug: No

U.S. FDA-regulated Device: No

U.S. FDA IND/IDE: No

Human Subjects Review: Board Status: Approved
Approval Number: 7.253.458
Board Name: Comitê de Ética e Pesquisa
Board Affiliation: University of Sao Paulo
Phone:

Address:

Rua Ovídio Pires de Campos, 225 - 5º andar - 55 11 26617585, Zip Code: 05.403-905

Data Monitoring: No

### Study Description

Brief Summary: This single-blind, randomized controlled trial investigates the effects of a remotely delivered intervention based on mental practice (MP) combined with physical practice (PP) on freezing of gait (FOG) severity in people with idiopathic Parkinson's disease (PD). Fifty participants will be randomly assigned to either the experimental group (MP + PP) or a control group (PP + stretching). The intervention consists of 10 video-supervised sessions over 6 weeks. Primary outcomes include the Rapid Turn Test and percentage of time spent with FOG (%FOG). Secondary outcomes include the New Freezing of Gait Questionnaire (NFOG-Q), Movement Disorders Society-Unified Parkinson's Disease Rating Scale (MDS-UPDRS), Parkinson's Disease Questionnaire-39 (PDQ-39), and Telephone Montreal Cognitive Assessment (T-MoCA). The study aims to demonstrate whether MP using Dynamic Neuro-Cognitive Imagery (DNI) techniques can effectively reduce FOG severity and improve motor and cognitive function in people with PD.

Detailed Description: This trial aims to evaluate a home-based remote intervention delivered via encrypted video calls by physiotherapists trained in both Mental Practice (MP) and Physical Practice (PP) protocols. Participants undergo 10 structured training sessions, each lasting 45–60 minutes, across six weeks. The experimental intervention consists of two blocks of first-person kinesthetic MP and two blocks of corresponding PP. The MP is guided using DNI principles, emphasizing goal-directed control, body schema reinforcement, and environmental context. Each block lasts approximately 10 minutes and focuses on gait-related challenges. Safety measures, including environmental checks and the presence of a caregiver (when applicable), are verified before each session. The control group performs the same physical practice but replaces the MP with guided seated stretching exercises. Assessments are conducted at baseline, post-intervention, and 30-day follow-up, by a blinded evaluator. The Rapid Turn Test is video-recorded and independently analyzed. A repeated measures ANOVA will be used for statistical analysis.

### Conditions

Conditions: Parkinson Disease
Freezing of Gait

Keywords: Parkinson disease
Freezing of Gait
Mental Practice
Telerehabilitation
Dynamic Neuro-Cognitive Imagery
Motor Imagery
Gait Disorders

### Study Design

Study Type: Interventional

Primary Purpose: Treatment

Study Phase: N/A

Interventional Study Model: Parallel Assignment
A single-blind, randomized controlled trial.

Number of Arms: 2

Masking: Single (Outcomes Assessor)
Outcome assessors blinded to group allocation.

Allocation: Randomized

Enrollment: 50 [Anticipated]

### Arms and Interventions

| Arms | Assigned Interventions |
| --- | --- |
| Experimental: Experimental Group (EG)<br>In the Experimental Group (EG), the intervention will consist of 10 remote sessions, each including two blocks of Mental Practice (MP) and two blocks of corresponding Physical Practice (PP), focuses on gait-related challenges. | Behavioral: Experimental intervention<br>Participants in this group will undergo 10 remote sessions over 6 weeks, each session lasting 45-60 minutes. Each session consists of: (1) two blocks of first-person kinesthetic MP, conducted while seated with eyes closed and guided using DNI principles; and (2) two blocks of corresponding PP, involving simulated gait components such as ankle mobility, postural weight shifts, and short-distance walking under real-time supervision. Each block lasts approximately 10 minutes. The MP emphasizes attentional focus, sensory detail, and task-specific goals. Sessions are conducted synchronously via video call by trained physiotherapists.<br>Other Names:<br>• (Mental Practice + Physical Practice) |
| Active Comparator: Control Group (CG)<br>In the Control Group (CG), the intervention will consist of 10 remote sessions, each including two blocks of seated upper-limb stretching exercises followed by two blocks of Physical Practice (PP), with the same structure and duration as in the Experimental Group (EG). | Behavioral: Control Intervention<br>Participants in the control group will receive 10 remote sessions over 6 weeks, each session lasting 45-60 minutes. Each session consists of the same PP as the experimental group but replaces motor imagery with two 10-minute seated stretching block, focusing on the upper limbs and trunk. The PP follows the same structure and duration as in the experimental group. All sessions are delivered remotely via live video by physiotherapists trained in both protocols. Volume, frequency, and interaction levels are matched between groups to ensure equivalence.<br>Other Names:<br>• (Physical Practice + Stretching) |

### Outcome Measures

Primary Outcome Measure:

1. Rapid Turns Test
   A provocative freezing test. The individuals will be instructed to spin around their own axis repeatedly, in both directions, at high speed. A Percentage of time with FOG (%FOG) will be performed based on video recordings of Rapid Turns Test to measure the actual severity of freezing, calculated using the formula (total duration of FOG during the test * 100) / total duration of the test.

   [Time Frame: Change from Baseline to immediately Post-intervention, and then at 30 days post-intervention (Follow-up).]

Secondary Outcome Measure:

2. New Freezing of Gait Questionnaire (NFOG-Q)
   A self reported questionnaire that assesses the clinical aspects of FOG and the impact on quality of life. The total scores range from 0 to 28, with higher scores indicating greater severity impact of FOG.

   [Time Frame: Change from Baseline to immediately Post-intervention, and then at 30 days post-intervention (Follow-up).]
3. Movement Disorders Society - Unified Parkinson's Disease Rating Scale (MDS-UPDRS) Part II
   It is a widely recognized scale used to evaluate and track the progression of Parkinson's Disease (PD). The scale includes 42 items, categorized into four sections: Part I (non-motor aspects of daily living), Part II (motor aspects of daily living), Part III (motor evaluation), and Part IV (motor complications). Scores range from 0 to 4, with lower scores indicating better functional status. In Part II, the evaluator asks the questions, and the responses are based on the individual's self-reported motor performance in daily activities.

   [Time Frame: Change from Baseline to immediately Post-intervention, and then at 30 days post-intervention .]
4. Parkinson Disease Questionnaire - 39 (PDQ-39)
   A questionnaire consisting of 39 questions distributed across eight domains (mobility - ten items; activities of daily living - six items; emotional well being - six items; social support - three items; physical discomfort - three items; stigma - four items; and communication - three items). Each item can be answered with one of five predetermined responses (never, rarely, sometimes, frequently, and always). The score for each item ranges from 0 to 4, with the total score ranging from 0 to 100, where the lowest score reflects better quality of life.

   [Time Frame: Change from Baseline to immediately Post-intervention, and then at 30 days post-intervention (follow-up).]
5. Telephone Montreal Cognitive Assessment (T-MoCA)
   The instrument evaluates six cognitive domains, including naming, memory, attention, language, abstraction, and orientation. The scale does not require the use of paper, pencil or visual stimuli and has a total score of 22 points, with higher scores indicating normal cognitive performance.

   [Time Frame: Change from Baseline to immediately Post-intervention, and then at 30 days post-intervention (follow-up).]

### Eligibility

Minimum Age:

Maximum Age:

Sex: All

Gender Based: No

Accepts Healthy Volunteers: No

Criteria: Inclusion Criteria:

- Clinical diagnosis of idiopathic PD;
- Use of dopaminergic medication;
- Experiencing FOG (positive response to the first question of the New Freezing of Gait Questionnaire - NFOG-Q);
- Able to walk independently at home;
- Access to internet and video call device;
- Agree to participate in the study.

Exclusion Criteria:

- Other neurological disorders;
- Severe cardiovascular and/or respiratory alterations;
- Uncorrected visual and/or auditory alterations;
- Cognitive impairment, detectable through the Telephone Montreal Cognitive Assessment (T-MoCA, < 12);

- Inability to perform motor imageryT during the administration of the Kinesthetic and Visual Imagery Questionnaire - 20 (KVIQ-20, < 20)

### Contacts/Locations

Central Contact Person: Maria Elisa P Piemonte, PT, PHD


Central Contact Backup: Paloma R Silva, PT


Study Officials: NOTE : Study Official is required by the WHO and ICMJE.

Locations: **Brazil**
University of Sao Paulo
[Recruiting]
Sao Paulo, Brazil, 05508-900
Contact: Maria Elisa P Piemonte, PT, PHD 55 11 30917451
Contact: Paloma R Silva, PT 55 11 976193193
Principal Investigator: Maria Elisa P Piemonte, PT, PHD
Principal Investigator: Paloma R Silva, PT

### IPDSharing

Plan to Share IPD:

### References

Citations:

Links:

Available IPD/Information:
